# Hand Dermatitis among Health Care Workers during the COVID-19 Pandemic: Prevalence, and Risk Factors

**DOI:** 10.1101/2022.06.27.22276976

**Authors:** Omolbanin Motamed-Rezaei, Gholamreza Sharif-Zadeh, Hadis Rajabipour, Farnaz Jahani, Hamed Lotfi

## Abstract

**Background:** Health care workers (HCWs) need to perform new preventive measures to protect themselves and patients against ongoing COVID-19 transmission, which can increase the occurrence of hand dermatitis (HD) among them.

**Objectives:** This study aimed to investigate the prevalence of HD among HCWs and its possible risk factors in IRAN.

**Methods:** A survey of 159 HCWS working in university hospitals was performed between August to September 2020. Research data were collected via the standardized Nordic Occupational Skin Questionnaire (NOSQ-2002).

**Results:** The prevalence of HD in the study population was 51.6%. Females had a 3.84 fold higher risk of HD than males (confidence interval (CI): 1.85-8). HCWs older than 40 years and those who aged 30-39 years had a 9.6 and 1.72 fold higher risk of HD than HCWs aged 20-29 years (CI: 2.6-35.7; CI: 0.87-3.4, respectively). A significant association was found between the prevalence of HD among HCWs and working hours per week, and wearing gloves (P<0.05).

**Conclusion:** Possible risk factors for developing HD among HCWs are female gender and older age. Preventive measures for HD are needed for HCWs, especially during the COVID- 19 pandemic.

## 1 Introduction

The Corona virus disease of 2019 (COVID-19) first appeared in December 2019 in Wuhan, China. Later, the World Health Organization (WHO) declared it a pandemic on March 11, 2020.^1^ The Center for Diseases Control (CDC) and World Health Organization (WHO) recommended preventive measures to hinder the spread of this novel virus (SARS-CoV-2). These measures emphasize the importance of social distancing, wearing a facemask and gloves, frequent hand washing with water and soap, or using alcohol-based hand sanitizers.^2-3^ These preventive measures for health care workers (HCWs), as a critical point of infection control, to protect themselves and prevent cross-transmission and spread of pathogens has already been emphasized.^4-5^ Although these recommendations may be effective against infectious diseases, they may alter integrity of the skin resulting in skin barrier dysfunction, and increased risk of hand dermatitis (HD).^6-7^

Contact dermatitis is a skin disorder that occurs when the skin encounters irritants or allergens. Contact dermatitis is the most common occupational disease, including irritant contact dermatitis (80%) and allergic contact dermatitis (20%).^8-9^ prolonged glove wearing and frequent hand hygiene using detergents, soaps, water, alcohol-based sanitizers, and other disinfectants are known risk factors for developing contact dermatitis. therefore, HCWs are at higher risk of developing contact dermatitis.^9^ Studies before the COVID-19 pandemic reported that prevalence of contact dermatitis affecting the hand of HCWs ranges from 12% to 50%^10-12^, Although, the reported prevalence of contact dermatitis in the studies from different countries during the COVID-19 outbreak have been increased.^13-16^

To date, there is limited data on the prevalence of HD among HCWs in Iran during the COVID-19 pandemic. We aimed to determine the prevalence of HD among HCWs in three university hospitals in Birjand, Iran, during the COVID-19 pandemic. Furthermore, we investigated the association between HD and other related factors.

## 2. Materials and Methods

### 2.1. Study design

The present study is a descriptive-analytic study conducted from August to September 2020 in Birjand, Iran. Cluster sampling was carried out in three university hospitals affiliated with Birjand University of Medical Sciences, including: Razi hospital, Valiasr hospital, and Imam Reza hospital. Birjand is the capital of South Khorasan province in eastern Iran. The protocol of study was approved by the Committee on Research Ethics at Birjand University of Medical Sciences with the approved No. 3789. (Ethic code IR.Bums.REC.1398.8).

### 2.2. Sample size calculation

A total sample size of 159 HCWs, who were defined as first-line workers performing patient-related duties, was calculated through the convenience sampling method. HCWs that have been working at the hospitals for at least one year were included in the study. Those who did not complete the questionnaire completely were excluded.

### 2.3. Data collection

Data was collected using a validated self-administered questionnaire. The history of occupational HD was determined using standardized Nordic Occupational Skin Questionnaire version 2002 (NOSQ-2002)^17^, which included two forms: The first form included questions about demographic information including age, gender, level of education, occupation, marital status, work experience, workplace ward, working hours per week, etc; the second form included questions about the most important occupational and environmental risk factors for formation of HD (i.e. history of allergy, history of HD, wearing gloves, frequency of hand washing, etc), and symptoms associated with HD (i.e. itching, burning, redness, dry skin with scaling, irritation, pain, and tiny water blister on the hand, wrist, or forearm) to determine prevalence of HD and associated factors among HCWs.^18^ In this questionnaire, having any of dermatitis-associated symptoms over the last six month (length of time HCWs were involved in caring for COVID-19) considered as dermatitis.

### 2.4. Data quality control

The use of this standardized questionnaire makes it possible to compare the results with other studies. The English version of NOSQ was translated to Persian by a professional translator familiar with medical terminology; thereafter, a panel of experts of academics (two dermatologists and eight faculty members of nursing and midwifery) familiar with the terminologies reviewed and confirmed the validity of the questionnaire. The reliability of the questionnaire was also confirmed by calculation of Cronbach’s α coefficient of 82%.^19^

### 2.5. Data analysis

Chi-square and Logistic regression tests were used for data comparisons, considering α level of 0.05. Data analysis was done using SPSS version 18. P-value < 0.05 was considered as significant threshold. Data were shown as mean ± SD and number (percent).

## 3. Results

The subjects were 159 HCWs. The participants’ mean age was 31.7 ± 10 years. The majority of the subjects participating in the study, 111 (66.9%) were female, and the highest number of HCWs, 129 (81.1%) were nurses. Demographic data of the HCWs participating in this study are shown in Table 1.

**Table 1.** Demographic data of the health care workers.

| Variable | Frequency | Percentage |
| --- | --- | --- |
| **Gender** |  |  |
| Male | 48 | 30.2 |
| Female | 111 | 69.8 |
| **Education Levels** |  |  |
| Diploma | 11 | 6.9 |
| Bachelor | 125 | 78.6 |
| MSc | 23 | 14.5 |
| **Duties** |  |  |
| Nurse | 129 | 81.1 |
| Auxiliary Nurse | 10 | 6.3 |
| Supervisor | 11 | 6.9 |
| Midwife | 5 | 3.1 |
| **Marital Status** |  |  |
| Single | 36 | 22.6 |
| Married | 123 | 77.4 |

Table 2 shows the prevalence of HD based on different variables of HCWs participating in this study. Eighty two (51.6%) of the study subjects had HD; the prevalence of HD had an almost equal distribution among HCWs. The prevalence of HD was significantly higher (P<0.05) in females (61.3%) than in males (29.2%). The prevalence of HD had an almost equal distribution among HCWs with a diploma (45.5%) and a bachelor (49.6%) educational degree. However, most of the HCWs with a MSc degree (65.2%) reported to have HD. The prevalence of HD was highest (86.4%) in the group older than 40 years and lowest (39.7%) in the 20-29 year group. Among 133 HCWs who currently use gloves, 66 (58.4%) had HD and the prevalence of HD was reported equal among those who did not wear gloves. The highest prevalence of HD (62.4%) was found in HCWs working fewer than 50 hours per week, and declined with increasing working hours.

**Table 2.** Prevalence of hand dermatitis based on different variables among study subjects

| Variable | Hand Dermatitis<br>Yes<br>Frequency<br>(Percentage) | Hand Dermatitis<br>No<br>Frequency<br>(Percentage) | Odds ratio<br>(OR) | Confidence<br>interval<br>(CI) | P value |
| --- | --- | --- | --- | --- | --- |
| **Total** | 82 (51.6) | 77 (48.4) |  |  |  |
| **Gender** |  |  |  |  |  |
| Male | 14 (29.2) | 34 (70.8) | 1 | 1.85 - 8 | P < 0.001 |
| Female | 68 (61.3) | 43 (38.7) | 3.84 |  |  |
| **Education Levels** |  |  |  |  |  |
| Diploma | 5 (45.5) | 6 (54.5) | 1 |  | P = 0.35 |
| Bachelor | 62 (49.6) | 63 (50.4) | 1.18 | 0.34 – 4.1 |  |
| MSc | 15 (65.2) | 8 (34.8) | 3.25 | 0.52 – 9.7 |  |
| **Age** |  |  |  |  |  |
| 20-29 years old | 29 (39.7) | 44 (60.3) | 1 |  | P = 0.001 |
| 30-39 years old | 34 (53.1) | 30 (46.9) | 1.72 | 0.87 – 3.4 |  |
| >40 years old | 19 (86.4) | 3 (13.6) | 9.6 | 2.6 – 35.7 |  |
| **Working hours** |  |  |  |  |  |
| <50 hours / week | 53 (62.4) | 38 (37.6) | 1 |  | P < 0.001 |
| ≥50 hours / week | 19 (32.8) | 39 (67.2) | 0.29 | 0.15 – 0.58 |  |
| **Wearing gloves** |  |  |  |  |  |
| Yes now | 66 (58.4) | 47 (41.6) | 1 |  | P = 0.021 |
| Yes but not now | 14 (33.3) | 28 (66.7) | 0.36 | 0.17 – 0.75 |  |
| Never | 2 (50) | 2 (50) | 0.71 | 0.96 – 5.2 |  |

Based on the results obtained from logistic regression test, female HCWs had a 3.84 fold increased risk of HD compared with males (confidence interval (CI): 1.85-8; P< 0.001). HCWs older than 40 years and those who aged 30-39 years had a 9.6 and 1.72 fold increased risk of HD compared with HCWs aged 20-29 years (CI: 2.6-35.7; CI: 0.87-3.4; P=0.001; P=0.001, respectively). Additionally, the prevalence of HD in HCWs who worked fewer than 50 hours per week was significantly higher than those who worked 50 hours or more per week (Odds Ratio (OR): 0.29; CI: 0.15-0.58; P< 0.001). Furthermore, results obtained from Chi-square test showed that the prevalence of HD among HCWs who are currently wearing gloves was significantly higher than those who never used gloves (OR: 0.71; CI: 0.96-5.2; P<0.05). Although, most of the HCWs with a MSc degree are reported to have HD, there was no significant association between prevalence of HD in HCWs and educational level (P>0.05).

## 4. Discussion

Hand dermatitis is a common occupational disease, with a prevalence of 12-50% among HCWs prior to the COVID-19 pandemic.^10-12^ in the current study the prevalence of HD was 51.6% among HCWs at university hospitals of Birjand, Iran, during the COVID-19 pandemic. Additionally, a significant correlation was found between the prevalence of HD and female gender, age, working hours per week, and wearing gloves. The results of our study is consistent with other studies conducted on skin damages among HCWs during the COVID-19 pandemic, which showed an increase in the prevalence of HD. Lan et al.^20^, in March 2020, reported the first evidence of increasing prevalence of skin damage among HCWs in China. It was reported that the prevalence of hand skin damage was 74.5%, which was probably related to frequent hand hygiene and wearing gloves for a longer time. A study conducted by Aydin et al.^13^, (2020) in Turkey, reported that the frequency of HD among nurses was 70.9%. Another study from that country conducted by Erdem et al.^16^, (2020) showed that the frequency of HD was 50.5% of the HCWs working in COVID-19 wards. Similarly, Imani Khanegah et al.^18^, (2020) in another province of Iran, reported that 65.7% of nurses working in COVID-19 ward in Ardabil Province had HD. Other studies from different countries reported the prevalence of symptoms associated with contact dermatitis of 46.4% in Saudi Arabia^15^, 90.4% in Germany^21^, 82.6% in Ireland^22^, 80% in Italy^23^, and 73.1% in China.^24^ The disparity in the prevalence is probably due to the different study design, population size, and genetic differences. Moreover, some studies assessed all types of skin damage, which reported overall HCWs’ skin damage during the COVID-19 pandemic, while in the current study we have just assessed the prevalence of HD. A study conducted by Tousi et al.^25^, (2006) in the same country as the present study, and reported that the prevalence of HD was 43.9% among HCWs in a university hospital of Tehran. Given these results, the prevalence of HD clearly increased among HCWs during the COVID-19 pandemic.

In the present study, we found a significantly higher prevalence of HD in females than in males. In line with our study, Celik et al.^26^, (2020) conducted a study on HCWs and showed that hand eczema was significantly higher in women. Likewise, Hamnerius et al.^27^, (2017) reported similar results either. Additionally, a large population-based Norwegian survey showed that the overall lifetime prevalence of HD was higher in women than men.^28^ The higher prevalence of dermatitis in women than men could be attributed to the role of women in housekeeping and more exposure to water, detergents, and Irritants.^26,29-30^ However, some studies reported that prevalence of HD was not associated with gender.^31-32^

The results of our study showed that there was a positive correlation between age of HCWs and the prevalence of HD, and the highest prevalence of HD was found in HCWs older than 40 years. Two studies conducted on Danish children reported that children with a higher age were at a higher risk of developing HD. They attributed their observations to a higher frequency of hand washing in children with higher age.^29,33-34^ On the other hand, the prevalence of HD was varied at different ages in other studies;^30,34-35^ therefore, age could not be considered as a direct risk factor for developing HD. This finding is probably due to the higher prevalence of HD in HCWs who have more years of working experience.^36^

In the current study, a negative correlation was found between the risk of developing HD and the working hours per week among HCWs. Lampel et al.^11^, found no significant association between the prevalence of HD in inpatient nurses working at an academic hospital and their working hours per week. By contrast, Özyazicioğlu et al. ^32^, reported that the incidence of HD in the fourth-year nursing students was significantly higher than the third-year students. They attributed this finding to the higher working hours of the fourth-year students compared with the third-year students, which was inconsistent with our findings. A possible speculative explanation for this could be increased exposure time of HCWs to other irritants at home while housekeeping, or the fact that HCWs with HD may work fewer hours per week.

There are some known risk factors for developing contact dermatitis including frequent hand washing, long term wearing gloves, and using disinfectants.^9^ It has been reported that SARS-CoV-2 could be transmitted through contact, thus proper hand hygiene is one of the main ways to prevent the COVID-19 spreading.^37^ Previous studies reported a significant increase in the frequency of hand washing and using disinfectants among HCWs during the COVID-19 pandemic.^21,38^ Repeated exposure to soap and water, and disinfectants could alter the integrity of the skin and damage protective effect of this organ by changing the PH of the epidermis, depleting the lipid barrier, and increasing trans-epidermal water loss, which finally leads to contact dermatitis. Paradoxically prolonged wearing of gloves may cause maceration and irritant contact dermatitis of the hand skin by over-hydrating the stratum corneum layer.^9^ Additionally, wearing latex gloves could develop skin reactions through immunoglobulin E-mediated hypersensitivity to latex, latex allergy, and irritant contact dermatitis.^39-40^ In the present study, 58.4% of HCWs who currently used gloves had HD. Similarly, Kaihui et al.^41^, (2020) reported that 88.5% of HCWs using long term of latex gloves during the COVID-19 outbreak complained adverse skin reactions.

HD is one of the main factors that can reduce the quality of life and productivity of HCWs, and may cause physical discomfort and increase their absenteeism either.^42-43^ Therefore, the results of our study may help prevent HD among HCWs, especially those who are at higher risk.

## 5. Conclusion

Although hand hygiene is an effective method to prevent transmission of infectious diseases such as COVID-19, it may increase the risk of hand dermatitis formation. Our results showed that the prevalence of HD was 51.6% among HCWs during the COVID-19 pandemic. Additionally, it was revealed that female gender, and older age might be possible risk factors for the occurrence of HD among HCWs. Therefore, this data can help hospitals to provide education on HD and acquaint HCWs with proper measures for prevention.

## Data Availability

All data produced in the present study are available upon reasonable request to the authors

## Authors Contributions

OM designed the study, supervised the project. GS analyzed the data and also performed statistical analysis. HR collected the research data. FJ collaborated on manuscript preparation and also contributed to interpretation of data. HL prepared the manuscript and critically revised the manuscript; besides, all authors read and approved the final version of the manuscript.

## Acknowledgement

We sincerely thank the help of the Vice-Chancellor for Research of Birjand University of Medical Sciences for the study financing.

## Conflicts of Interest

None.

## Ethical Approval and Consent to Participate

This research was approved by the research ethics committee on Research Ethics at Birjand University of Medical Sciences with the approved code of 3789 (ethics code ir.bums.REC.1398.8).

## Data Availability

The datasets generated and analyzed during the current study are available from the corresponding author on reasonable request.

## Funding

None.

## Supplementary Data

The supplementary data are available on the journal’s website and the published article.

## Notes

### Competing Interest Statement

The authors have declared no competing interest.

### Funding Statement

This study did not receive any funding

### Author Declarations

the research ethics committee on Research Ethics at Birjand University of Medical Sciences approved code: 3789 ethics code: ir.bums.REC.1398.8

## References

1. Coronavirus disease 2019 (COVID-19) Situation Report-101 2020 Available at: https://www.who.int/docs/default-source/coronaviruse/situation-reports/20200430-sitrep-101-COVID-19pdf?sfvrsn=2ba4e093_2. Accessed July 10,2020.

2. Centers for Disease Control & Prevention. Coronavirus Disease 2019 (COVID-19). June 2021. http://www.cdc.gov/coronavirus/2019-ncov/prevent-getting-sick/hand-sanitizer.Html. Last accessed June 25, 2021.

3. WHO. Coronavirus Disease (COVID-19) advice for the public. https://www.who.int/emergencies/diseases/novel-coronavirus-2019/advicefor-public. 2020.

4. Pires D, Pittet D. Hand hygiene mantra: teach, monitor, improve, and celebrate. J Hosp Infect. 2017 Apr;95(4):335–337. doi: 10.1016/j.Jhin.2017.03.009.

5. Verbeek JH, Rajamaki B, Ijaz S, Tikka C, Ruotsalainen JH, Edmond MB, Sauni R, Kilinc Balci FS. Personal protective equipment for preventing highly infectious diseases due to exposure to contaminated body fluids in healthcare staff. Cochrane Database Syst Rev. 2019 Jul 1;7(7):Cd011621. doi: 10.1002/14651858.Cd011621.Pub3. Update in: Cochrane Database Syst Rev. 2020 Apr 15;4:Cd011621.

6. Dejonckheere G, Herman A, Baeck M. Allergic contact dermatitis caused by synthetic rubber gloves in healthcare workers: Sensitization to 1,3-diphenylguanidine is common. Contact Dermatitis. 2019 Sep;81(3):167–173. doi: 10.1111/cod.13269.

7. Afshar M, Lotfi H, Hassanzadeh Taheri MM, Zardast M. The Effect of Aqueous-Alcoholic Extract of Toothbrush Tree (Salvadora Persica) on the healing of Second-degree Skin Burns in BALB/c Mice. Pharm Sci. 2022. doi:10.34172/ps.2022.1.

8. Dhingra N, Shemer A, Correa da Rosa J, Rozenblit M, Fuentes-Duculan J, Gittler JK, Finney R, Czarnowicki T, Zheng X, Xu H, Estrada YD, Cardinale I, Suárez-Fariñas M, Krueger JG, Guttman-assky e. Molecular profiling of contact dermatitis skin identifies allergen-dependent differences in immune response. J Allergy Clin Immunol. 2014 Aug;134(2):362–72. doi: 10.1016/j.Jaci.2014.03.009.

9. Abdali S, Yu J. Occupational Dermatoses Related to Personal Protective Equipment Used During the COVID-19 Pandemic. Dermatol Clin. 2021 Oct;39(4):555–568. doi: 10.1016/j.Det.2021.05.009.

10. Ibler KS, Jemec GB, Flyvholm MA, Diepgen TL, Jensen A, Agner T. Hand eczema: Prevalence and risk factors of hand eczema in a population of 2274 healthcare workers. Contact Dermatitis. 2012 Oct;67(4):200–7. doi: 10.1111/j.1600-0536.2012.02105.x.

11. Lampel HP, Patel N, Boyse K, O’brien SH, Zirwas MJ. Prevalence of hand dermatitis in inpatient nurses at a united states hospital. Dermatitis. 2007 Sep;18(3):140–2. Doi: 10.2310/6620.2007.06024.

12. Van der meer EW, Boot CR, van der Gulden JW, Jungbauer FH, Coenraads PJ, Anema JR. Hand eczema among healthcare professionals in the Netherlands: Prevalence, absenteeism, and presenteeism. Contact Dermatitis. 2013 Sep;69(3):164–71. Doi: 10.1111/cod.12099.

13. Aydın Aİ, Atak M, Özyazıcıoğlu N, Dalkızan V. Hand Dermatitis among Nurses during the COVID-19 Pandemic: Frequency and Factors. Adv skin wound care. 2021 Dec 1;34(12):651–655. doi: 10.1097/01.Asw.0000765916.20726.41.

14. Lin P, Zhu S, Huang Y, Li L, Tao J, Lei T, Song J, Liu D, Chen L, Shi Y, Jiang S, Liu Q, Xie J, Chen H, Duan Y, Xia Y, Zhou Y, Mei Y, Zhou X, Wu J, Fang M, Meng Z, Li H. Adverse skin reactions among healthcare workers during the coronavirus disease 2019 outbreak: A survey in Wuhan and its surrounding regions. Br J Dermatol. 2020 Jul;183(1):190–192. doi: 10.1111/bjd.19089.

15. Alluhayyan OB, Alshahri BK, Farhat AM, Alsugair S, Siddiqui JJ, Alghabawy K, AlQefari GB, Alolayan WO, Abu Hashem IA. Occupational-Related Contact Dermatitis: Prevalence and Risk Factors Among Healthcare Workers in the Al’Qassim Region, Saudi Arabia During the COVID-19 Pandemic. Cureus. 2020 Oct 15;12(10):E10975. doi: 10.7759/cureus.10975.

16. Erdem Y, Altunay IK, Aksu Çerman A, Inal S, Ugurer E, Sivaz O, Kaya HE, Gulsunay IE, Sekerlisoy G, Vural O, Özkaya E. The risk of hand eczema in healthcare workers during the COVID-19 pandemic: Do we need specific attention or prevention strategies? Contact Dermatitis. 2020 Nov;83(5):422–423. doi: 10.1111/cod.13632.

17. Susitaival P, Flyvholm MA, Meding B, Kanerva L, Lindberg M, Svensson A, Olafsson JH. Nordic Occupational Skin Questionnaire (NOSQ-2002): a new tool for surveying occupational skin diseases and exposure. Contact Dermatitis. 2003 Aug;49(2):70–6. doi: 10.1111/j.0105-1873.2003.00159.x.

18. Imani Khanegah N, Ayadi N, Heidarzadeh M, Ajri-Khameslou M, Davari M. Investigating hand dermatitis among nurses in iran during the outbreak of COVID-19: Comparison of COVID and non-COVID wards. Npt. 2021;8(4):257–264.

19. Tamene A. Occupational Contact Dermatitis in Employees of Large-Scale Narcotic Crop Farms of Ethiopia: Prevalence and Risk Factors. A Self-Reported Study Using the Nordic Occupational Skin Questionnaire. Environ Health Insights. 2021 Oct 7;15:11786302211048378. doi: 10.1177/11786302211048378.

20. Lan J, Song Z, Miao X, Li H, Li Y, Dong L, Yang J, An X, Zhang Y, Yang L, Zhou N, Yang L, Li J, Cao J, Wang J, Tao J. Skin damage among health care workers managing coronavirus disease-2019. J Am Acad Dermatol. 2020 May;82(5):1215–1216. doi: 10.1016/j.Jaad.2020.03.014.

21. Guertler A, Moellhoff N, Schenck TL, Hagen CS, Kendziora B, Giunta RE, French LE, Reinholz M. Onset of occupational hand eczema among healthcare workers during the SARS-CoV-2 pandemic: Comparing a single surgical site with a COVID-19 intensive care unit. Contact Dermatitis. 2020 Aug;83(2):108–114. doi: 10.1111/cod.13618.

22. Kiely LF, Moloney E, O’sullivan G, Eustace JA, Gallagher J, Bourke JF. Irritant contact dermatitis in healthcare workers as a result of the COVID-19 pandemic: a cross-sectional study. Clin Exp Dermatol. 2021 Jan;46(1):142–144. doi: 10.1111/ced.14397.

23. Rizzi A, Inchingolo R, Viola M, Boldrini L, Lenkowicz J, Lohmeyer FM, De Simone FM, Staiti D, Sarnari C, Gasbarrini A, Nucera E. Occupational hand dermatitis web survey in a university hospital during COVID-19 pandemic: The SHIELD study. Med Lav. 2021 Aug 26;112(4):320–326. doi: 10.23749/mdl.V112i4.11670.

24. Pei S, Xue Y, Zhao S, Alexander N, Mohamad G, Chen X, Yin M. Occupational skin conditions on the front line: A survey among 484 Chinese healthcare professionals caring for Covid-19 patients. J Eur Acad Dermatol Venereol. 2020 Aug;34(8):E354–e357. doi: 10.1111/jdv.16570.

25. Tousi P, Rahmati M, Taheri A. Hand Dermatitis Among Staff in Loghman Hospital in Tehran. Pajoohande. 2008; 12 (6) :521–526 [persian]..

26. Celik V, Ozkars MY. An overlooked risk for healthcare workers amid COVID-19: Occupational hand eczema. North Clin Istanb. 2020 Nov 18;7(6):527–533. doi: 10.14744/nci.2020.45722.

27. Hamnerius N, Svedman C, Bergendorff O, Björk J, Bruze M, Pontén A. Wet work exposure and hand eczema among healthcare workers: a cross-sectional study. Br J Dermatol. 2018 Feb;178(2):452–461. doi: 10.1111/bjd.15813.

28. Vindenes HK, Svanes C, Lygre SHL, Hollund BE, Langhammer A, Bertelsen RJ. Prevalence of, and work-related risk factors for, hand eczema in a Norwegian general population (The Hunt Study). Contact Dermatitis. 2017 Oct;77(4):214–223. doi: 10.1111/cod.12800.

29. Simonsen AB, Ruge IF, Quaade AS, Johansen jJD, Thyssen JP, Zachariae C. Increased occurrence of hand eczema in young children following the Danish hand hygiene recommendations during the COVID-19 pandemic. Contact Dermatitis. 2021 mar;84(3):144–152. doi: 10.1111/cod.13727.

30. Simonsen AB, Ruge If, Quaade AS, Johansen JD, Thyssen JP, Zachariae C. High incidence of hand eczema in Danish schoolchildren following intensive hand hygiene during the COVID-19 pandemic: a nationwide questionnaire study. Br J Dermatol. 2020 Nov;183(5):975–976. doi: 10.1111/bjd.19413.

31. Zhang D, Zhang J, Sun S, Gao M, Tong A. Prevalence and risk factors of hand eczema in hospital-based nurses in northern China. Australas J Dermatol. 2018 Aug;5(3):E194–e197. doi: 10.1111/ajd.12672.

32. Özyazicioğlu N, Sürenler S, Aydin Aİ, Atak M. Hand Dermatitis in Nursing Students. Adv Skin Wound Care. 2020 Apr;33(4):213–216. doi: 10.1097/01.Asw.0000655472.02780.E0.

33. Borch L, Thorsteinsson K, Warner TC, Mikkelsen CS, Bjerring P, Lundbye-Christensen S, Arvesen K, Hagstroem S. COVID-19 reopening causes high risk of irritant contact dermatitis in children. Dan Med J. 2020 Aug 6;67(9):A05200357.

34. Drewitz KP, Stark KJ, Zimmermann ME, Heid IM, Apfelbacher CJ. Frequency of hand eczema in the elderly: Cross-sectional findings from the German AugUr study. Contact Dermatitis. 2021 Nov;85(5):489–493. doi: 10.1111/cod.13920.

35. Voorberg AN, Loman L, Schuttelaar MLA. Prevalence and Severity of Hand Eczema in the Dutch General Population: A Cross-sectional, Questionnaire Study within the Lifelines Cohort Study. Acta Derm Venereol. 2022 Jan 5;102:Adv00626. doi: 10.2340/actadv.V101.432.

36. Lan CC, Feng WW, Lu YW, Wu CS, Hung ST, Hsu HY, Yu HS, Ko YC, Lee CH, Yang YH, Chen GS. Hand eczema among university hospital nursing staff: Identification of high-risk sector and impact on quality of life. Contact Dermatitis. 2008 Nov;59(5):301–6. doi: 10.1111/j.1600-0536.2008.01439.x.

37. Wu YC, Chen CS, Chan YJ. The outbreak of COVID-19: An overview. J Chin Med Assoc. 2020 Mar;83(3):217–220. doi: 10.1097/jcma.0000000000000270.

38. Erdem Y, Inal S, Sivaz O, Copur S, Boluk KN, Ugurer E, Kaya HE, Gulsunay IE, Sekerlisoy G, Vural O, Altunay IK, Aksu Çerman A, Özkaya E. How does working in pandemic units affect the risk of occupational hand eczema in healthcare workers during the coronavirus disease-2019 (COVID-19) pandemic: A comparative analysis with nonpandemic units. Contact Dermatitis. 2021 Apr 1:10.1111/cod.13853. doi: 10.1111/cod.13853.

39. Douglas R, Morton J, Czarny D, O’hehir RE. Prevalence of IgE-mediated allergy to latex in hospital nursing staff. Aust N Z J Med. 1997 Apr;27(2):165–9. doi: 10.1111/j.1445-5994.1997.Tb00933.x.

40. Weido AJ, Sim TC. The burgeoning problem of latex sensitivity. Surgical gloves are only the beginning. Postgrad Med. 1995 Sep;98(3):173–4, 179-82, 184.

41. Hu K, Fan J, Li X, Gou X, Li X, Zhou X. The adverse skin reactions of health care workers using personal protective equipment for COVID-19. Medicine (Baltimore). 2020 Jun 12;99(24):e20603. doi: 10.1097/md.0000000000020603.

42. Mekonnen TH, Yenealem DG, Tolosa BM. Self-report occupational-related contact dermatitis: Prevalence and risk factors among healthcare workers in Gondar town, Northwest Ethiopia, 2018-a cross-sectional study. Environ Health Prev Med. 2019 Feb 14;24(1):11. doi: 10.1186/s12199-019-0765-0.

43. Yüksel YT, Nørreslet LB, Flachs EM, Ebbehøj NE, Agner T. Hand eczema, wet work exposure, and quality of life in health care workers in Denmark during the COVID-19 pandemic. JAAD Int. 2022 Jun;7:86–94. doi: 10.1016/j.Jdin.2022.02.009.

